# Household transmissions of SARS-CoV-2 in the time of unprecedented travel lockdown in China

**DOI:** 10.1101/2020.03.02.20029868

**Authors:** Xiao-Ke Xu, Xiao-Fan Liu, Lin Wang, Sheikh Taslim Ali, Zhanwei Du, Paolo Bosetti, Benjamin J. Cowling, Ye Wu

## Abstract

**Importance:** Severe acute respiratory syndrome coronavirus 2 (SARS-CoV-2) emerged in the city of Wuhan, China, in December 2019 and then spread globally. Limited information is available for characterizing epidemiological features and transmission patterns in the regions outside of Hubei Province. Detailed data on transmission at the individual level could be an asset to understand the transmission mechanisms and respective patterns in different settings.

**Objective:** To reconstruct infection events and transmission clusters of SARS-CoV-2 for estimating epidemiological characteristics at household and non-household settings, including super-spreading events, serial intervals, age- and gender-stratified risks of infection in China outside of Hubei Province.

**Design, Setting, and Participants:** 9,120 confirmed cases reported online by 264 Chinese urban Health Commissions in 27 provinces from January 20 to February 19, 2020. A line-list database is established with detailed information on demographic, social and epidemiological characteristics. The infection events are categorized into the household and non-household settings.

**Exposures:** Confirmed cases of SARS-CoV-2 infections.

**Main Outcomes and Measures:** Information about demographic characteristics, social relationships, travel history, timelines of potential exposure, symptom onset, confirmation, and hospitalization were extracted from online public reports. 1,407 infection events formed 643 transmission clusters were reconstructed.

**Results:** In total 34 primary cases were identified as super spreaders, and 5 household super-spreading events were observed. The mean serial interval is estimated to be 4.95 days (standard deviation: 5.24 days) and 5.19 days (standard deviation: 5.28 days) for households and non-household transmissions, respectively. The risk of being infected outside of households is higher for age groups between 18 and 64 years, whereas the hazard of being infected within households is higher for age groups of young (<18) and elderly (>65) people.

**Conclusions and Relevance:** The identification of super-spreading events, short serial intervals, and a higher risk of being infected outside of households for male people of age between 18 and 64 indicate a significant barrier to the case identification and management, which calls for intensive non-pharmaceutical interventions (e.g. cancellation of public gathering, limited access of public services) as the potential mitigation strategies.

**Key Points:** *Question:* What epidemiological characteristics and risk factors are associated with household and non-household transmissions of the severe acute respiratory syndrome coronavirus 2 (SARS-CoV-2) in China outside of Hubei Province?

*Findings:* In this epidemiological study analyzing 1,407 SARS-CoV-2 infection events reported between 20 January 2020 and 19 February 2020, 643 transmission clusters were reconstructed to demonstrate the non-negligible frequency of super-spreading events, short duration of serial intervals, and a higher risk of being infected outside of household for male people of age between 18 and 64 years.

*Meaning:* These findings provide epidemiological features and risk estimates for both household and non-household transmissions of SARS-CoV-2 in China outside of Hubei Province.

## Introduction

In December 2019, a novel coronavirus (severe acute respiratory syndrome coronavirus 2, SARS-CoV-2) emerged in Wuhan city of Hubei Province in China. World Health Organization (WHO) announced a public health emergency of international significance on 30 January 2020 ^1^ and classified the threat as a global pandemic on 11 March 2020 ^2^. More than 118,326 confirmed cases and 4,292 deaths have been reported as of 11 March 2020. The majority of cases in China (67,773) were from Hubei Province^3^.

On 23 January 2020, China raised the national emergency response to the highest level, triggered an unprecedented travel ban starting from the lockdown of Wuhan on 23 January, and 14 cities nearby Wuhan on 24 January, and more than 30 provinces thereafter (Figure 1a). Although this countrywide travel lockdown was aimed to interrupt case exportations from the epicenter, we have estimated 130 (95% CrI: 190, 369) high-risk cities that have introduced SARS-CoV-2 cases prior to Wuhan’s lockdown ^4^. Recent studies^5,6^ report similar estimates for the rapid geographic expansion of SARS-CoV-2. These coincide with the frequent reporting of infection events in China outside of Hubei Province during the three weeks following Wuhan’s lockdown (Figure 1c).

**Figure 1:**
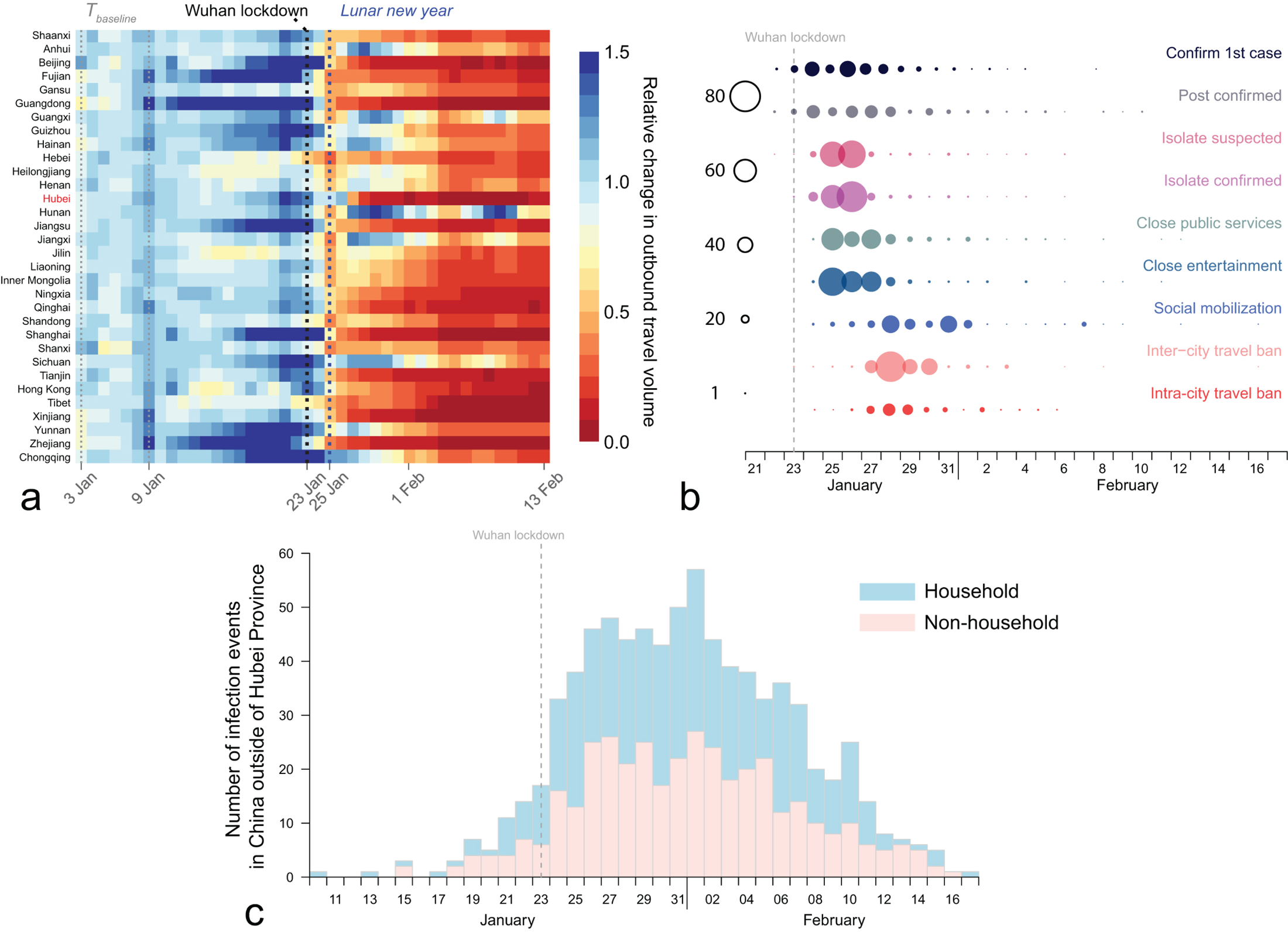
Changing patterns of population movements, non-pharmaceutical control measures, and epidemic curve of infection events in China outside of Hubei Province. (a) Relative change in the outbound travel volume (i.e., daily number of passengers) moving out of each province in China from 1 January to 13 February 2020. The baseline daily outbound travel volume of a given province is estimated by the mean daily number of passengers moving out of that province between 3 January 2020 and 9 January 2020 (i.e., the last week prior to the start of the Spring Festival travel season). The relative change in outbound travel volume of province *i* on a given date *T*_*i*_ is measured by the ratio between the estimated number of passengers moving out of province *i* on date *T*_*i*_ and the baseline daily outbound travel volume of province *i*. Each row denotes a province, with Hubei Province highlighted in red. Columns from left to right are ordered by calendar days. For the color bar, the dark red and blue indicate a significant decrease and increase in the outbound travel volume, respectively. The Spring Festival travel season officially started on 10 January 2020. The Lunar New Year holiday started on 25 January 2020 and was extended to 8 February 2020. The lockdown of Wuhan started on 23 January 2020. The end date of Spring Festival travel season is not clear because of the countrywide travel lockdown. (b) The timing of epidemiological events and implementation of non-pharmaceutical control measures in 263 cities in China. The size of circles is proportional to the number of cities. The vertical grey dashed line indicates the timing of Wuhan’s lockdown. (c) The date of symptom onset for secondary cases caused by household (light pink) and non-household (light blue) primary infections.

Since the last week of January 2020, more than 260 Chinese cities have implemented intensive non-pharmaceutical controls (Figure 1b), which brought the epidemic under control ^7,8,9^. However, as of 19 February 2020, this epidemic still has caused >10,000 cases in China outside of Hubei Province. To enhance public health preparedness and public awareness, Chinese health authorities have publicly reported detailed records of confirmed cases since 20 January 2020. This provides a unique resource and an opportunity for understanding the transmission patterns, routes, and risk factors of the COVID-19 epidemic.

## Methods

### Data Collection

In China, 27 provincial and 264 urban health commissions have publicly posted 9,120 confirmed case reports online since 20 January 2020, which comprises 72.7% of all cases confirmed in China outside of Hubei Province by 19 February (Table S9). We compiled a line-list database from these reports with extracted demographic characteristics, social relationships, travel history, timelines of potential exposure, symptom onset, confirmation and hospitalization. We obtained the daily population movement data in China between 1 January and 13 February 2020 from Baidu Qianxi Web Portal (https://qianxi.baidu.com/). We estimated the daily number of passengers leaving each province by using the migration scale index for moving out of that province each day. More details on our real-time mobility and line-list case data are provided in Supplemental Material and will be available at our GitHub.

### Statistical Analysis

We identified 1,407 infection events with known social relationships. For each infection event, we term the infector the *primary* case and the infectee the *secondary* case. We also consider connected chains of confirmed cases; we term the original case the *index* and the entire chain of cases, including the index, the *transmission cluster* (Figure 2a).

**Figure 2:**
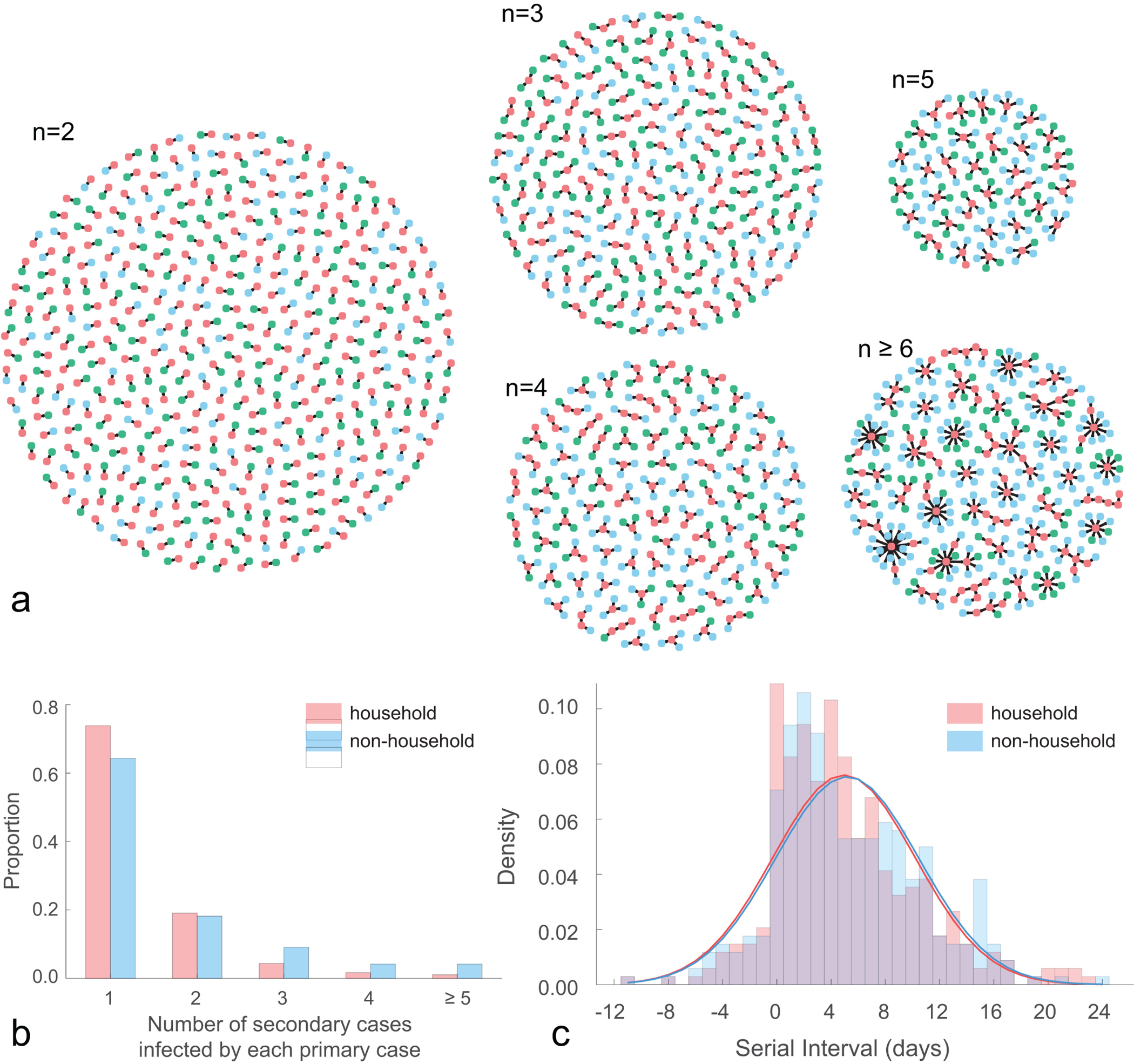
COVID-19 transmission clusters. (a) 643 transmission clusters, stratified by the size of cluster *n*. Red, green and blue nodes denote primary cases, household secondary cases and non-household secondary cases, respectively. (b) Distribution of the number of secondary infections caused by each of the 809 primary cases. (c) Distribution of serial intervals, which is fitted to 679 infection events with known symptom onset timelines. For household transmissions, fitting with a normal distribution suggests a mean serial interval of 4.95 days (standard deviation: 5.24 days); for non-household transmissions, fitting with a normal distribution suggests a mean serial interval of 5.19 days (standard deviation: 5.28 days). Alternative distributions fitted for the serial interval can be found in Table S5.

We stratify infection events by household versus non-household transmissions, where household includes any infection event among members within the same family (e.g., between parents and children), and non-household include all others (e.g. colleagues, classmates). The numbers of household (662) and non-household (745) infection events are almost even.

For each infection event with known symptom onset timelines, we compute the serial interval, i.e., the number of days between the reported symptom onset date for the primary case and that for the secondary case. We estimate the distribution of serial intervals by fitting a normal distribution to 679 infection events with known serial intervals (Supplemental Material).

The age-stratified hazard of infection for household relative to non-household contacts is estimated by the ratio between the probability that a secondary case of age group *b* was infected by a primary case of age group *a* within the same household and the probability that a secondary case of age group *b* was infected by a primary case of age group *a* outside of households. Gender-specific hazard of infection is measured similarly (Supplemental Material).

## Results

We reconstructed 643 transmission clusters from 1,407 infection events (Figure 2a). The sizes of 587 transmission clusters are smaller than 5, whereas the size of the largest cluster exceeds 20. We observed 34 primary cases acting as super spreaders. Stratification by household shows that 356, 92, and 34 primary cases infected only 1 member, 2 members, and at least 3 familial members within households, respectively; 276, 78, and 75 primary cases infected 1, 2, and at least 3 secondary cases outside households, respectively (Figure 2b). Only 5 household super-spreading events were observed.

The mean serial interval is estimated to be 4.95 days (standard deviation: 5.24 days) within households, and 5.19 days (standard deviation: 5.28 days) for non-household transmissions. Notably, 26 of 339 household- and 29 of 340 non-household infection events reported negative-valued serial intervals, implying pre-symptomatic transmission.

The hazard of being infected within households is higher for age groups of young (<18) and elderly (>65) people, whereas the hazard of being infected outside of households is higher for age groups between 18 and 64 years (Table 1a). Primary cases of elderly (>65) people are more prone to cause household infections. Hazard of infection between different genders is higher for households than non-household transmission (Table 1b).

**Table 1:**
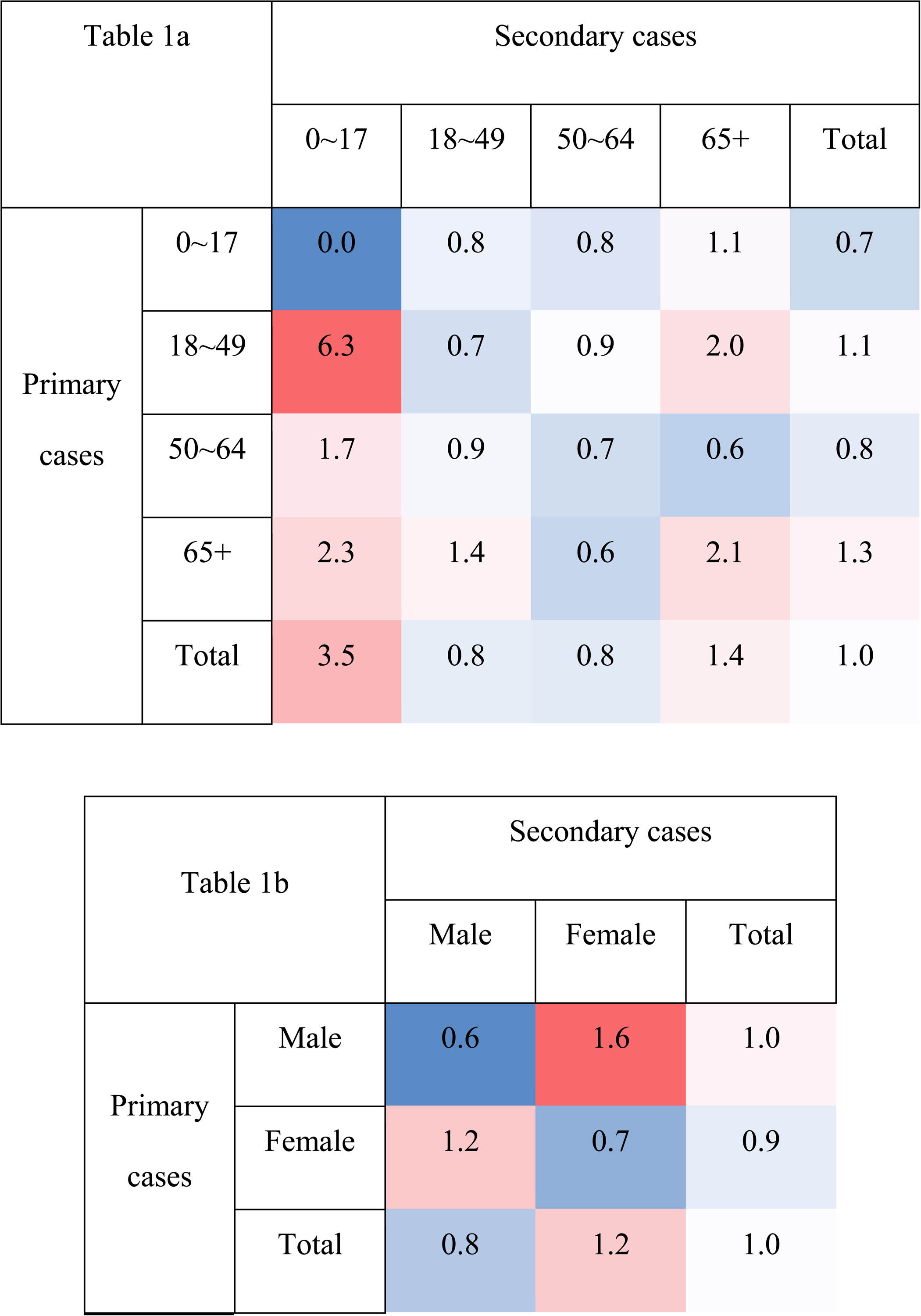
Hazard of infection stratified by age or gender. (a) Age-stratified hazard of infection for the household relative to non-household transmission. (b) Gender-specific hazard of infection for the household relative to non-household transmission. Red or blue shades indicate an increased or decreased hazard of infection within households relative to outside of households, respectively.

## Discussion

We reconstructed 1,407 infection events formed 643 transmission clusters from 9,120 COVID-19 cases confirmed in China outside of Hubei Province as of 19 February 2020. The entire database will be available at our GitHub.

Although super-spreading events have been reported by media reports and a recent study ^11^ which focuses on special settings (e.g., restaurant, chalet) before February 2020, systematic analyses with a sufficiently large sample size is lacking. Here, we identified 34 primary cases classified as super-spreader. Majority of the super-spreading events involve more non-household transmissions than household transmissions. This indicates the importance of non-pharmaceutical interventions (e.g. cancellation of public gatherings, limited access of public services) as the potential mitigation strategies (see Table S2).

Household study helps identify risk factors for certain demographic groups ^12^. The age-stratified and gender-specific hazard of infection suggests a higher risk of infection within households for age groups of young (<18) and elderly (>65) and female people. The higher risk of being infected outside of households for male people of age between 18 and 64 years indicates their role in driving household secondary infections in the households included in our data, perhaps because these were travelers from Wuhan of working age.

We identified 55 infection events (∼4%) with the secondary case reporting symptom onset prior to the primary case, which is consistent with our preliminary analysis ^4^ and recent clinical reports ^13,14^. Negative-valued serial intervals indicate the potential occurrence of pre-symptomatic transmissions. We estimate that the mean serial interval is ∼5 days for both household and non-household infections, which is considerably shorter than the mean serial interval estimated for SARS (e.g., 8.4 days ^15^) and MERS (e.g. 7.6 days ^16^). These evidences impose a significant barrier to case identification and management.

Our findings have several limitations. First, the size of each household and the primary cases without secondary infections are unknown from original disclosures. This may give biased estimates if we estimate the household reproduction number and secondary attack rate from raw data. Field surveys will be helpful to adjust biases. Second, the information on nosocomial infections is unknown from original disclosures, so that the observation of super-spreading events may be less common from our dataset.

## Data Availability

The data will be available for download after the manuscript is published on a journal.

## Acknowledgements

We thank Simon Cauchemez, Lauren Ancel Meyers,Henrik Salje, Juliette Paireau, Dongsheng Luo, and Lanfang Hu for helpful discussions.

